# Syndromic detectability of haemorrhagic fever outbreaks

**DOI:** 10.1101/2020.03.28.20019463

**Authors:** Emma E. Glennon, Freya L. Jephcott, Alexandra Oti, Colin J. Carlson, Fausto A. Bustos Carillo, C. Reed Hranac, Edyth Parker, James L. N. Wood, Olivier Restif

## Abstract

Late detection of emerging viral transmission allows outbreaks to spread uncontrolled, the devastating consequences of which are exemplified by recent epidemics of Ebola virus disease. Especially challenging in places with sparse healthcare, limited diagnostic capacity, and public health infrastructure, syndromes with overlapping febrile presentations easily evade early detection. There is a clear need for evidence-based and context-dependent tools to make syndromic surveillance more efficient. Using published data on symptom presentation and incidence of 21 febrile syndromes, we develop a novel algorithm for aetiological identification of case clusters and demonstrate its ability to identify outbreaks of dengue, malaria, typhoid fever, and meningococcal disease based on clinical data from past outbreaks. We then apply the same algorithm to simulated outbreaks to systematically estimate the syndromic detectability of outbreaks of all 21 syndromes. We show that while most rare haemorrhagic fevers are clinically distinct from most endemic fevers in sub-Saharan Africa, VHF detectability is limited even under conditions of perfect syndromic surveillance. Furthermore, even large clusters (20+ cases) of filoviral diseases cannot be routinely distinguished by the clinical criteria present in their case definitions alone; we show that simple syndromic case definitions are insensitive to rare fevers across most of the region. We map the estimated detectability of Ebola virus disease across sub-Saharan Africa, based on geospatially mapped estimates of malaria, dengue, and other fevers with overlapping syndromes. We demonstrate “hidden hotspots” where Ebola virus is likely to spill over from wildlife and also transmit undetected for many cases. Such places may represent both the locations of past unobserved outbreaks and potential future origins for larger epidemics. Finally, we consider the implications of these results for improved locally relevant syndromic surveillance and the consequences of syndemics and under-resourced health infrastructure for infectious disease emergence.

## Introduction

Outbreaks of Ebola virus disease (EVD) and other emerging zoonotic diseases are most easily contained during the first few transmission cycles, when cases are localised and small in number ^1^. Numbers of case contacts—and corresponding difficulties and costs of containment—grow exponentially in an epidemic’s earliest stages and may quickly overwhelm local healthcare capacity ^2^. Despite the critical importance of detection in these early stages, we have rarely detected EVD soon enough; we recently estimated that only 25-30% of past EVD outbreaks of five cases or fewer have been detected and reported ^3^. Many other emerging diseases, including other viral haemorrhagic fevers (VHFs), have received less attention than EVD, and detection rates are likely to be even lower in the early stages of an epidemic ^4^.

Prompt detection of emerging viruses is limited by access to health care, local diagnostic capacity, and reporting efficiency. Rural clinics in West Africa, for example, generally lack local access to laboratory methods for definitive confirmatory testing for pathogens such as yellow fever and dengue viruses, and are even less likely to have tests for Ebola virus and similarly rare pathogens ^5,6^. In this context, early detection of VHF outbreaks requires that clinicians differentially diagnose often-nonspecific febrile syndromes against a background of widespread febrile illness, a particularly challenging process in places with limited health and public health infrastructure ^6–8^. Even in the relatively well-financed international responses to Ebola outbreaks, triaging patients for Ebola risk based on clinical presentation is a difficult task ^9–11^. Furthermore, practitioners’ knowledge of clinical profiles of rare or emerging diseases may be inaccurate or inappropriate for many settings, especially for vector-borne diseases caused by a variety of strains and for which mild cases are often unobserved ^12,13^.

In addition to infrastructure for detection, the endemic context in which spillover occurs contributes to the detectability of diseases with similar presentations. In resource-limited clinical settings, treating as many patients as possible is often a higher priority than diagnosing any specific disease or performing particular surveillance functions. Diagnosis is often performed through trial and error with treatment rather than through the use of laboratory testing ^5^. In parts of sub-Saharan Africa, for example, febrile illnesses are often misdiagnosed as common diseases such as malaria, even in the presence of unusual clinical features or negative malaria test results ^14^. Furthermore, these settings face compounding challenges caused by limited training and infrastructure. Healthcare workers are not well positioned to be able to quickly and thoroughly interpret relatively rare and/or subjective clinical features such as hiccups or fatigue—and under-resourced places with limited diagnostic capacity are also likely to face more endemic noise from other poorly controlled diseases ^5,15^.

Until diagnostic capacity and public health infrastructure improve in many low- and middle-income countries, syndromic surveillance is likely to remain the primary method of detecting emerging outbreaks. New methods are needed to improve the efficacy of this approach. To model the current state of syndromic surveillance and understand opportunities for improvement, we develop a Bayesian algorithm for aetiological identification of outbreaks based on case clinical features. We test this algorithm against data from 87 outbreaks of dengue, meningococcal disease, malaria, and typhoid fever. We then simulate outbreaks of 21 different syndromes to estimate the detectability of each (i.e., the estimated probability of a cluster of cases being correctly identified by a syndromic surveillance regime, based on its clinical features) as a function of the number of cases. We examine spatial heterogeneity in syndrome detectability across sub-Saharan Africa by incorporating estimates of malaria, dengue, yellow fever, and diarrheal disease incidence at high spatial resolution, as well as the ecological risk distributions of Crimean-Congo haemorrhagic fever and Ebola virus disease. By estimating detectability across this spatially variant endemic context, we identify potential “hidden hotspots” where VHFs are especially likely to spill over and transmit among people.

## Materials and methods

### Data collection

We selected 21 potentially haemorrhagic febrile syndromes based on their clinical presentations and incidences (see Text S1 for inclusion criteria). These VHFs span a range of ecological and epidemiological characteristics, including mammalian zoonoses (e.g., EVD, Marburg virus disease) and vector-borne diseases (e.g., epidemic typhus and Crimean-Congo haemorrhagic fever). We also generated profiles for the more common febrile syndromes that comprise the endemic context of selected VHFs. Based on an extensive search of the clinical literature, including case and outbreak reports, we then estimated the probability distributions of a range of signs and symptoms for each syndrome (i.e., clinical features; see Text S1). We also gathered estimates of the incidence of each disease by country from the Global Burden of Disease Survey; where these were not available for a disease, we estimated spillover rates from the literature. We incorporated and performed a sensitivity analysis on the effects of a “spillover scalar” to weight the prior probabilities of these rarer diseases in the algorithm (see Text S1).

### Syndromic identification and detectability

To summarise overlap in clinical features among the 21 syndromes, we created a matrix of syndromic distances, with all signs and symptoms normalized by the incidence-weighted probability of clinical feature occurrence among all cases. We defined the syndromic distance between syndromes as the Euclidean distance across each pair of sign/symptom probabilities, weighted by a scalar derived from t-SNE ^16^ to account for collinearity between symptoms (*θ*_*k*_; Text S2).

For each syndrome, we then calculated normalised probabilities (*P*_*c,n*_) of *c* cases of syndrome *n* being identified as any of *M* total syndromes. Where clinical features *x* are simulated from beta-binomial draws (*x*_*k,n*_ *= X∼Bin(c, p∼Beta(v*_*k*_*p*_*k*.*n*_, *v*_*k*_ (1 *-p*_*k*.*n*_))), *I*_*m*_ is the incidence of syndrome *m*, and *v*_*k*_ is estimated to approximate heterogeneity in presentation for each clinical feature (Text S1), these values are:

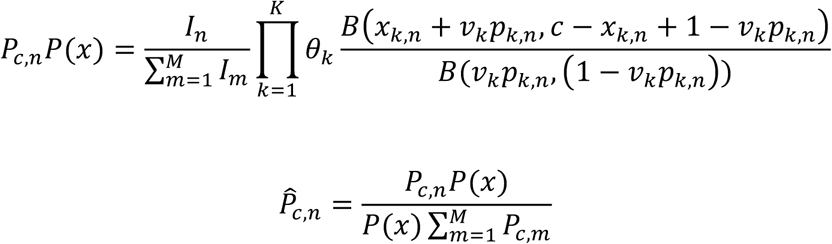

We refer to the quantity 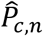 as the ‘detectability’ of syndrome *n* at cluster size *c*, representing the theoretical maximum sensitivity of syndromic surveillance of syndrome clusters based on their expected clinical presentations (considering only the *K=*18 clinical features for which we collected data). The expected detectable size of a syndrome *n* within its local disease context is the minimum cluster size *c* for which 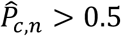. Unless otherwise stated, we used the medians (and appropriate quantiles for 95% confidence intervals and interquartile ranges) of 100 Monte Carlo simulations as detectability estimates.

### Analysis

We first calibrated the spillover scalar and collinearity parameters using to a grid search (Text S3). To understand model performance on real data, we applied the aetiological identification algorithm to a published dataset of clinical feature data from 87 outbreaks of malaria, dengue, meningococcal disease, and typhoid fever in South Asia (Text S3). As the outbreaks in the dataset varied in terms of the number of clinical feature probabilities reported, we estimated both the probability of correct aetiological identification of the outbreak and the expected detectability of the outbreak given the clinical features reported.

We then estimated the clinical syndromic distances between each of the 21 syndromes and their detectability over the course of an outbreak (up to 20 cases) under different syndromic surveillance regimes (i.e., considering all symptoms, typical symptoms, or minimal VHF symptoms; Text S1). Finally, to estimate geospatial variability in the difficulty of detecting Ebola virus outbreaks, we estimated mean detectability of 5 simulated 10-case EVD clusters across sub-Saharan Africa using high-spatial resolution incidence estimates for malaria ^17,18^, yellow fever ^19^, dengue ^20^, Crimean-Congo haemorrhagic fever ^21^, diarrhoeal diseases, typhoid, and invasive non-typhoidal *Salmonella* ^22,23^ (Text S4).

## Results

### Algorithm performance on historical outbreaks

On the test set of 87 malaria, dengue, meningococcal septicaemia, and typhoid fever outbreaks from South Asia, the calibrated algorithm correctly identified the true outbreak cause in almost all cases, its accuracy increasing with the number of clinical features available and the size of the outbreak/symptom cluster (Fig. 1). Posterior probabilities of the true outbreak aetiologies, across all outbreaks and cluster sizes, were higher than the respective incidence-based prior probabilities in 94% of cases (Fig. S6-A). To account for variability in data quality—e.g., we expected poor algorithm performance when applied to those outbreaks for which only one or two clinical features were reported—we also estimated a performance ratio for each outbreak (Text S5). According to this ratio, malaria detection probabilities were consistently higher than expected, while dengue and meningococcal detection probabilities were consistent with expectations (Fig. S6-B). Algorithm performance on typhoid outbreaks was the least accurate, with seven of ten outbreaks predicted to have less than 10% of their expected probability (Fig. S6-B). Typhoid outbreaks with several clinical features reported tended to be misidentified as diarrhoeal diseases with a higher-than-expected probability, and those with few symptoms reported (in several instances, only a low probability of death and/or haemorrhage) tended to be misidentified as lower or upper respiratory infections with a higher-than-expected probability (Fig. S6-C).

**Figure 1.**
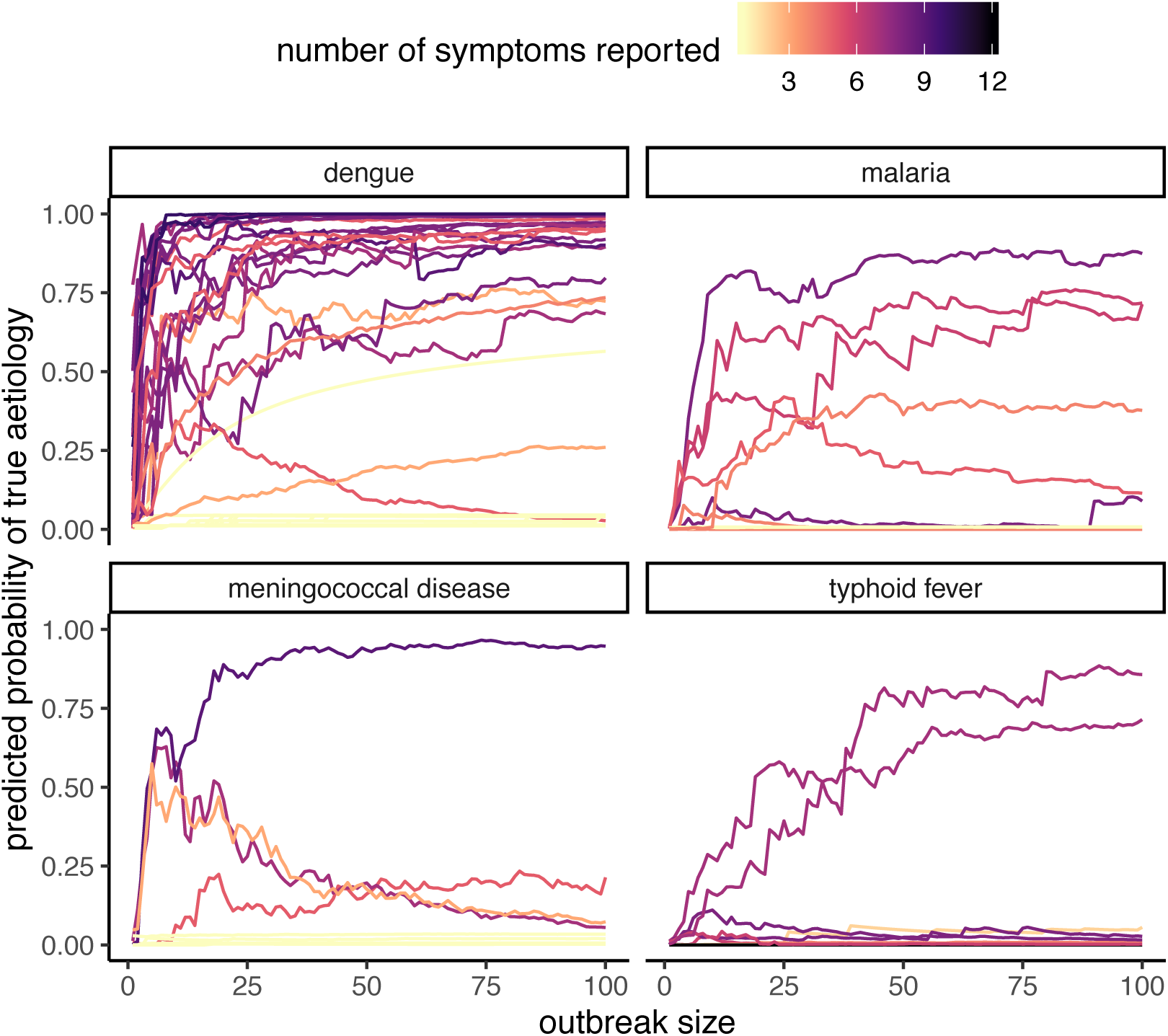
Posterior probabilities of correct algorithmic identification of dengue, malaria, meningococcal disease, and typhoid fever outbreaks in South Asia, as a function of the size of the simulated outbreak and the number of relevant clinical features reported for the outbreak. Each line corresponds to one real-world disease outbreak in South Asia, with a different set of reported clinical features available for each outbreak.

### Estimated detectability of febrile outbreaks

By applying our aetiological identification algorithm to simulated outbreaks of all 21 syndromes, we estimate the detectable sizes of each syndrome as well as likely misidentifications between syndromes (Fig. 2). We estimate, for example, that Ebola virus disease and Marburg virus disease (MVD) are most likely to be misidentified as dengue haemorrhagic fever, yellow fever, or typhoid fever. The clinical features caused by EVD and MVD outbreaks are more likely to be caused by other diseases until at least 6 and 7 cases occur, respectively. Before this detectability threshold, the balance of probabilities suggests a few cases of more common aetiologies are presenting with rare clinical features; after this threshold, the presentation of cases is unusual enough to suggest a common presentation of extremely rare aetiologies (i.e., filoviral disease). We summarise the estimated detectabilities—i.e., probabilities of correct aetiological identification—for each syndrome in Fig. 3, as a function of both cluster size and clinical features considered.

**Figure 2.**
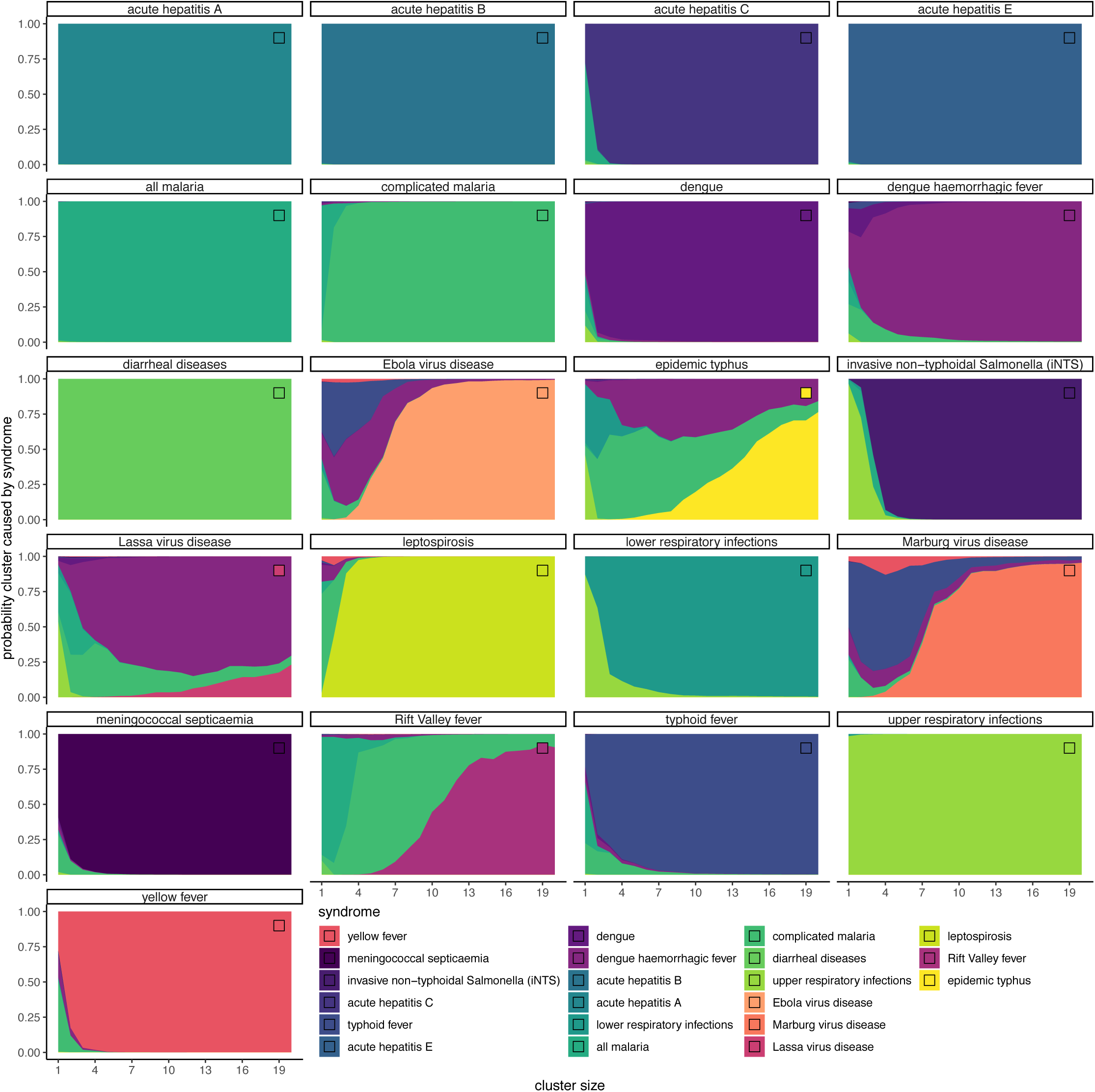
Aetiological posterior probabilities assigned to clinical feature clusters of between 1 and 20 cases, based on application of the aetiological identification algorithm to 100 simulated clusters of each syndrome, including all 18 clinical features. Squares in the upper right corner of each plot indicate the colour of the “true” syndrome, i.e., the syndrome used to simulate clinical feature clusters.

**Figure 3.**
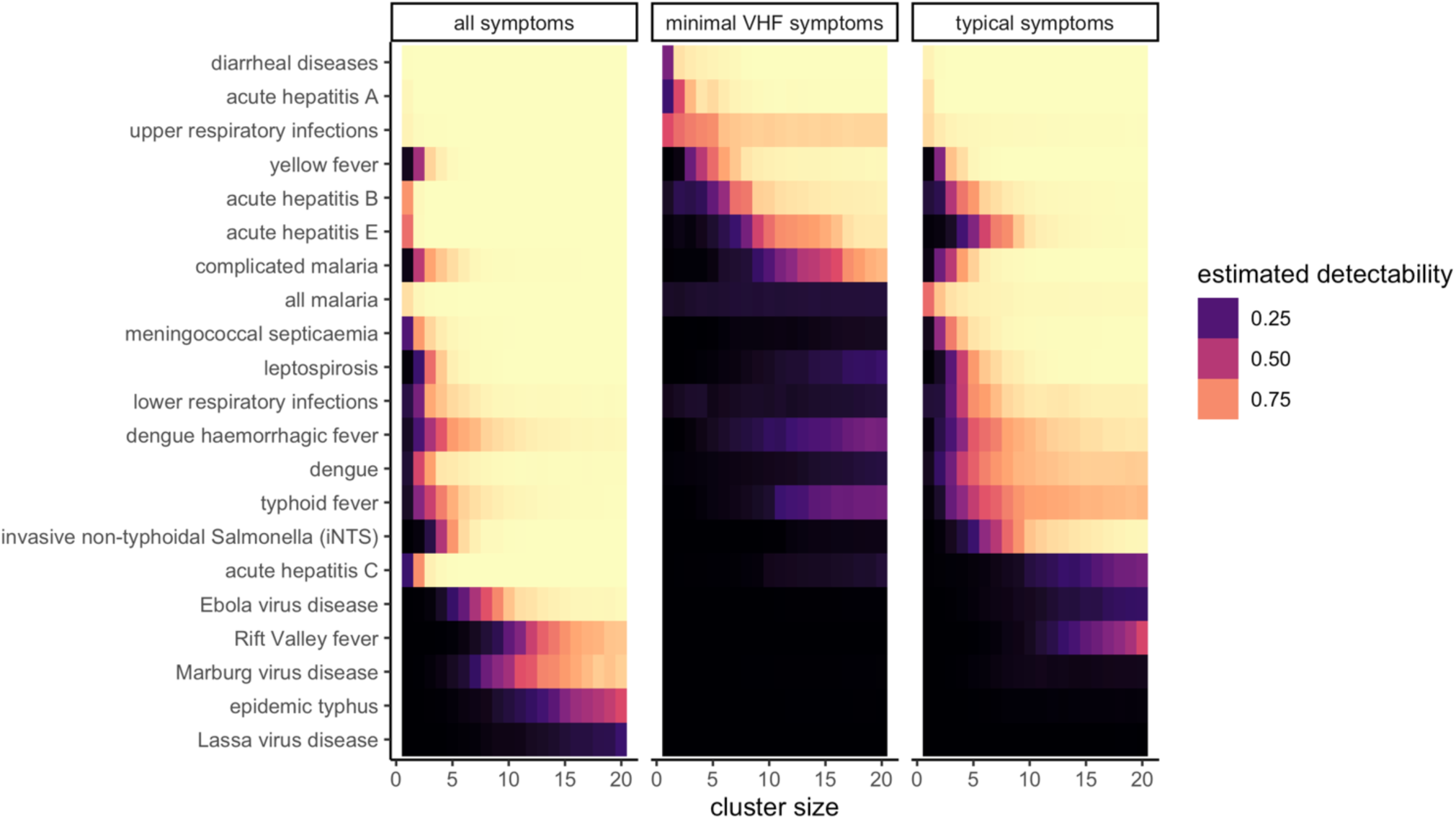
Detectability of selected infectious syndromes in sub-Saharan Africa when considering different sets of clinical feature. Left: all clinical features included in our database (i.e., fever, diarrhoea, death, fatigue or weakness, anorexia, nausea and/or vomiting, abdominal pain, any haemorrhage or bleeding, sore throat, cough, hiccups, headache, and jaundice). Centre: minimal viral haemorrhagic fever signs and symptoms (i.e., fever, death, hiccups, jaundice, and haemorrhage/bleeding). Right: only symptoms typical of a syndrome (i.e., those more common for a syndrome than for the incidence-weighted mean of all included syndromes).

Other rare VHFs and zoonotic diseases have greater syndromic overlap with malaria, including Rift Valley fever and epidemic typhus (Fig. 2); these two syndromes are not 50% detectable until clusters of 12 and 15 cases, respectively (Fig. 3). Lassa fever overlaps most with malaria at small cluster sizes (approx. 1-5 cases) and dengue haemorrhagic fever at larger sizes (approx. 5+ cases) and is not 50% detectable until a cluster size of 25 cases. Considering all 18 clinical features included in our database, most other syndromes reach 50% detectability within the first 5 cases; this includes moderately rare syndromes such as leptospirosis, yellow fever, and typhoid fever. Reducing the range of clinical features considered dramatically reduces identifiability of most syndromes (Fig 3). Considering only minimal VHF features (i.e., fever, haemorrhage/bleeding, death, hiccups, and jaundice) renders most syndromes undetectable by 20 cases, suggesting that e.g., case definitions for EVD or MVD are insufficient to detect them within the first 20 cases. However, the addition of nonstandard symptoms improves the detectability of filoviral diseases and other VHFs substantially.

### Spatial variation in detectability of Ebola virus disease outbreaks

Considering endemic context at a higher spatial resolution reveals heterogeneity in detectability and regions where EVD is most able to spread undetected (Fig. 4-A). Where existing risk maps for EVD consider the ecological niche of Ebola virus in reservoir hosts and/or human population density and movement, we add an entirely new layer to the EVD map representing variation in outbreak detectability. Considering detectability of EVD clusters in the context of existing geospatial estimates of spillover risk demonstrates potential “hidden hotspots” where EVD is both most able to spread undetected by syndromic surveillance and most likely to spill over from wildlife into people (Fig. 4-B). These potential hidden hotspots include central coastal West Africa, northern provinces of the Democratic Republic of the Congo, and the region joining Equatorial Guinea, Gabon, and Cameroon (Fig. 4-B).

**Figure 4.**
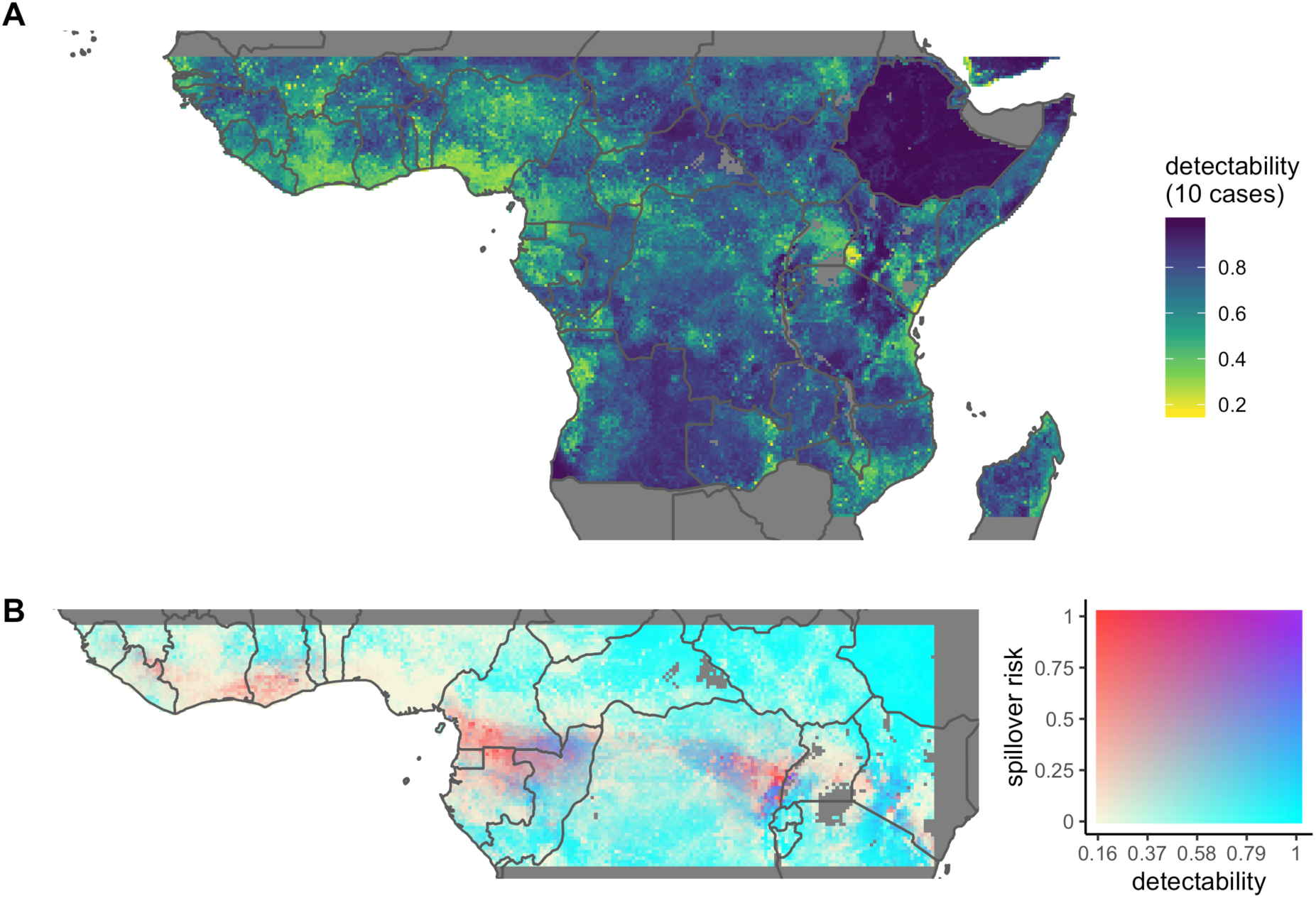
A. Estimated detectability of a cluster of ten cases of Ebola virus disease, based on full reporting of clinical features and incorporating geospatial estimates of per capita malaria, dengue, yellow fever, typhoid, invasive non-typhoidal *Salmonella*, Crimean-Congo-haemorrhagic fever, and diarrheal disease incidences. B. Estimated detectability of ten cases of EVD (blue) vs. expected spillover risk (red) ^24^.

## Discussion

Detection of case clusters is critical to breaking the chain of events that leads from a single zoonotic spillover event to a large-scale epidemic. Although the epidemiological process underlying this chain of events has received growing attention ^25^, the process of clinical detection has rarely been studied directly due to the inherent difficulties of measuring cases and outbreaks that were never recorded. Here we have attempted to address this gap by quantifying how the endemic context in which a disease occurs obscures syndromic signals.

We have introduced new measures of syndromic distance and a new model of syndromic surveillance and tested this model against a range of outbreaks with varying data quality, demonstrating its strong utility in identifying outbreak aetiologies. We have used these new methods to estimate the sensitivity of syndromic surveillance to a range of potentially haemorrhagic fevers. We have also quantified how syndromic overlap between febrile syndromes in sub-Saharan Africa affects the expected number of cases required to detect outbreaks across the region. Finally, by incorporating geospatial estimates of the incidences of common febrile syndromes, we have provided evidence for “hidden hotspots” where Ebola virus is both likely to spill over and especially difficult to detect, creating spaces suitable for unchecked transmission before detection and intervention is possible ^1^.

### Algorithmic outbreak identification and syndromic detectability

Our syndromic surveillance model, which combines the prior incidences and clinical profiles of a range of diseases to predict the causes of clinical feature clusters, presents a more mechanistic approach than other models intended for outbreak attribution ^26^. Among the primary limitations of expanding our approach is the time- and labour-intensive nature of parameterisation based on literature. Testing the aetiological identification algorithm against 87 real outbreaks highlights both the strengths of our approach and these data limitations; while we were frequently able to predict the true aetiology from limited dengue and malaria outbreak data, performance was less accurate for typhoid fever. Given more outbreak data— in terms of quantity, syndromes represented, and consistency of clinical feature reporting— we expect to be able to improve the sensitivity, specificity, and accuracy of the algorithm for categorising real outbreak data. As is, however, our mechanistic approach still enables strong insights into the relationships among outbreak detection, case definitions and the values of clinical features for disease identification, and local endemic context.

A central challenge to developing this approach has been the lack of standardised clinical data, especially given the high dimensionality of our model. For example, we have been unable to gather comparable data on non-communicable diseases or toxic aetiologies. Although haemorrhagic and febrile syndromes are unlikely to be misdiagnosed as non-infectious syndromes ^5^, the paucity of data on such syndromes may prevent such an approach from being used on, for example, syndromes presenting with jaundice or neurological symptoms. Particularly scant data is also available for many uncommon or seemingly insignificant clinical features, such as hiccups or headache. Although these features are mild, we nonetheless find that they are important for both the sensitivity and specificity of syndromic surveillance approaches. Additionally, few studies describing the presentation of syndromes describe heterogeneity in their presentation. We accounted for this heterogeneity by introducing wider clinical feature distributions for those syndromes which represent pooled aetiological agents (e.g., ascribing high heterogeneity to “diarrhoeal diseases,” which includes a range of pathogens, and moderate heterogeneity to most clinical features of EVD, which may be caused by several distinct filovirus species). However, more consistent and comprehensive reporting of clinical feature prevalence across strains and settings would enable stronger accounting for heterogeneity in disease presentation.

More broadly, using literature-derived clinical feature probabilities makes our model extremely high-dimensional, making thorough validation of and sensitivity analysis on all parameters prohibitively complex without standardised clinical datasets. However, all clinical feature occurrence parameters are clinically interpretable and approximate existing processes of expert clinical diagnosis. Our literature-derived parameter set can therefore be easily interpreted and refined, e.g., by expert clinicians, Bayesian or machine learning methods, or incorporation of standardised clinical data.

This parameter set therefore represents an important starting point to harnessing complex clinical information to identify outbreaks and undetected disease more quickly and accurately than current approaches. Our methods have the additional strengths of being scalable, clinically interpretable, tolerant of missing data, and useful in the presence of different clinical feature combinations. This flexibility renders our methods more practical for disease detection at a population level than, for example, the decision tree-based or machine learning methods commonly used by individual-level aetiological prediction models ^27–29^.

Furthermore, an advantage of our approach is its ability to exploit—rather than work against—the uncertainty inherent in syndromic data from diseases with heterogeneous presentations. Critically, this allows detection of diseases at population levels even before any individual-level diagnosis occurs (e.g., before the development of tests for rare/novel pathogens or in settings with insufficient diagnostic capacity).

Our approach has the potential to clarify aspects of disease emergence far beyond those presented here for viral haemorrhagic fevers. For example, upon expanding the range of signs/symptoms, diseases, and spatial data, it may be possible to estimate the properties of diseases least likely to be detectable by current surveillance infrastructure. The inclusion of more pathognomonic signs and symptoms in particular could improve our ability to distinguish between diseases. It would also be possible to estimate observation probabilities and predicted spillover locations of other poorly observed emerging diseases and to develop geographically specific case definitions. For example, application of similar methods to respiratory syndromic profiles could illuminate the factors affecting detection of SARS coronavirus 2 disease (COVID-19) outbreaks and rapidly adapt syndromic screening protocols to local contexts ^30^.

### Spatial detectability of Ebola virus disease

We demonstrate that filoviral diseases, along with yellow fever, represent a distinct and distinguishable group of syndromes based on their estimated syndromic distances from other febrile syndromes in sub-Saharan Africa. However, filoviral haemorrhagic fevers remain difficult to detect based on symptoms alone due to their relative rarity. A cluster of haemorrhagic fever with a high risk of mortality, for example, even with a low proportion of jaundice, is more likely to be caused by yellow fever than either Marburg virus disease or Ebola virus disease until well more than 20 cases have occurred. Consideration of every clinical feature in our dataset—the collection of which would require extensive health system coordination, universal health access, and deep clinical expertise—still only allows for attribution of EVD or MVD by the sixth and seventh case, respectively. In an area unaccustomed to such diseases or without adequate resources, months of transmission could occur silently, such as those which complicated control of the two largest Ebola outbreaks to date ^31–34^.

This analysis suggests that common clinical case definitions for filovirus diseases are insufficient for detection of viral haemorrhagic fever outbreaks, including EVD outbreaks, in many parts of West and Central Africa. Clusters of clinical features caused by an EVD outbreak, even if they are identified as related, are too commonly attributable to other febrile symptoms. Correctly identifying EVD cases based on case definitions alone is a known challenge even for Ebola treatment centres (ETCs) during ongoing outbreaks; addition of more specific clinical features including conjunctivitis and diarrhoea has been shown to improve the accuracy of ETC triage protocols ^11^. Correctly identifying the first cases of an outbreak—i.e., when filoviral disease is unlikely to be a common diagnostic consideration— requires even greater sensitivity and specificity to Ebola’s clinical manifestations than ETC triage. Furthermore, the variation we demonstrate in the detectability of EVD clusters across West and Central Africa indicates that appropriate case definitions for routine surveillance might require tailoring to local or regional endemic context.

Although a strength of our approach is the use of endemic disease data to understand rare and poorly observed diseases, it is still limited by the availability of appropriate ecological and epidemiological data. The relatively high predicted detectability of EVD outbreaks in Ethiopia, for example, may be an artefact of underestimated incidences of dengue, malaria, and yellow fever in the country (Fig. S7). More broadly, niche maps for filoviral and other zoonotic diseases with wildlife origins rely on incompletely observed human/primate outbreak data for validation ^35^. The Ebola virus spillover map used to generate Figure 4-B, for example, was fit to known instances of spillover in people (either directly from the putative reservoir or via intermediate hosts such as apes and duikers) ^24^. While cross-validated on subsets of known spillovers and built on independent, underlying ecological data (i.e., fruit bat and non-molossid microbat diversity, bat demographic patterns, and human population density), such models still face fundamental challenges related to potential spatially heterogeneous observation biases. More consistent ecological data collection, especially related to bat abundance estimates and migratory patterns, could help overcome these limitations and identify regions of high ecological spillover risk. Furthermore, it may be possible to understand the hidden hotspots we identify more deeply by integrating ecological and observational uncertainties into a single spillover modelling framework.

While niche maps for vector-borne diseases are somewhat more reliable, especially given careful surveillance and mapping of the *Aedes aegypti* mosquito ^36^, rarer and newer diseases are still less understood. The global dengue virus map has been refined over several years with an evidence-based, consensus-building process ^20,37,38^, and approximates risk well, even though the four circulating serotypes create a complex underlying pattern of immunity that complicates predictions of infection risk and outbreak severity ^39^. The yellow fever virus map for Africa is comparatively newer, and model uncertainty is still high in East and Central Africa, where occurrence data is comparatively sparse ^19^. Minor differences in niche model parameterization can produce major downstream differences in model-based inference ^35,40^; future work can capture this uncertainty by combining risk maps from several sources.

Similarly, future work may benefit from using models that are spatially tailored to the area of interest ^35^, or incorporate other dimensions of spatiotemporal heterogeneity like seasonal spillover risk. These dimensions might be especially informative for ruling out arboviral diseases ^41,42^, but could also be informative for filoviruses as spillover drivers become clearer^43^.

Syndemic interactions among diseases further complicate the detection process. Ecological risk factors are likely to be clustered within populations, especially for the handful of diseases that share zoonotic reservoirs (EVD and MVD) or mosquito vectors (dengue, yellow fever, and nearly a dozen other emerging viruses). Corresponding social risk factors—like food insecurity and water storage, or limited access to primary healthcare—are also highly spatially correlated, further increasing the odds that these diseases cluster together at fine spatial scales. These factors compound any potential biological associations between diseases ^44^, and resulting co-morbidities can alter clinical presentations, complicating both diagnosis and treatment. This makes outbreak dynamics more complex, and potentially more severe, than each disease would normally cause in isolation ^15^. This is a particular problem for emerging viruses, which may have poorly characterized clinical presentations and case definitions for several years after emergence ^45^; failure to characterise new syndromes in full can delay a national or global response by weeks or months, with strong consequences for the rate of epidemic spread ^1^. For example, recent work suggests that the majority of Zika cases in the Americas were likely misidentified as chikungunya and dengue, long before the outbreak was formally reported ^46^. Recent work also suggests that official case definitions for Zika by the World Health Organization miss a majority of paediatric Zika cases because the case definitions target the disease’s adult manifestation, which is substantially different than its appearance in children ^13^. Similar risks are apparent for EVD and MVD, especially in the hotspots we identify.

While some of our predicted hidden hotspots overlap with recently estimated hotspots for EVD epidemic risk, we introduce several new predicted areas of risk based on the difficulty of detection. For example, both southeastern Guinea and northeastern Democratic Republic of Congo emerged as hidden hotspots in our model, were identified as hotspots in recent epidemic risk models ^47,48^, and have served as the geographic origins of the two largest recorded Ebola epidemics. We also, however, predict EVD detection to be especially difficult in coastal West Africa—especially southern Ghana and Cote d’Ivoire—and in Equatorial Guinea, eastern Cameroon, and northern Gabon/Republic of Congo.

These regions may represent locations of historically unobserved outbreaks ^3^. These hidden hotspots represent only those places where Ebola virus disease is most likely to be undetectable by syndromic surveillance; they do not indicate anything else about the state of infrastructure for surveillance, public health coordination, or diagnosis. However, our predictions are supported by their correspondence with serosurveys and a lack of reported outbreaks despite high ecological suitability ^49–51^. Moderately high seroprevalence of filovirus antibodies has been observed in wildlife and/or human populations in these regions ^49,51–58^, but human outbreaks have rarely been reported ^59^. Reanalysis of historical outbreaks in these regions, especially those with ambiguous syndromic presentations, could provide further evidence about the nature of local ecology and surveillance.

### Implications for syndromic surveillance

In addition to understanding the broader ecology and epidemiology of emerging haemorrhagic fevers, our results suggest several opportunities for improving syndromic surveillance as currently practiced. Similar analyses could, for example, lead to the establishment of geographically adaptable case definitions that are maximally sensitive and specific to their local endemic contexts. Case definitions for emerging diseases are often variable over space and time even in well-observed outbreaks ^60^, and our methodology offers a starting point for consistent and quantitative validation of case definitions’ sensitivity and specificity in different contexts. Furthermore, although it seems unlikely that a healthcare worker would initiate the testing for diseases as rare as EVD or MVD outside the context of an on-going outbreak, the findings from our study do suggest a diagnostic testing algorithm of sorts. For instance, having all samples from suspected yellow fever cases that have tested negative for yellow fever automatically tested for EVD and MVD, could improve detection capacity even given diagnostic constraints at the healthcare facility-level. The data limitations encountered in our study highlight the need for standardised clinical data collection, especially for signs and symptoms rarely associated with the syndromes under consideration. Beyond improving syndromic models, such standardisation—and rapid sharing of data where appropriate ^61^—could enable algorithmic detection of outbreaks of misdiagnosed diseases based on key clinical metrics. This method could enhance and complement existing genomic ^62–65^ and statistical ^66–68^ approaches to outbreak detection.

Our results suggest that the best way to find filovirus outbreaks early is to build systems for controlling the endemic diseases that obscure them. Just as under-resourcing health systems creates multiple interacting effects that allow disease to thrive, improving basic public health infrastructure can have far-reaching effects. We demonstrate the benefits of diagnostic and surveillance infrastructure for improving detection of emerging haemorrhagic fevers, but also that supporting population health—including universal health coverage ^69,70^, water and sanitation infrastructure ^71,72^, and strategies to address the socio-political determinants of health ^73,74^—has compounding benefits in terms of disease detection. By reducing endemic burdens, for example, progress in these areas can make rare epidemics easier to detect (an indirect effect analogous to that of vaccination and sanitation for reducing antibiotic overprescription and antimicrobial resistance ^75^). Furthermore, the presence of well-equipped healthcare facilities and local public health officials are likely to enable reporting and control measures, reducing the risk of outbreaks spreading once started. Additional research within the framework we introduce could quantify these potential effects. Such holistic, system-wide approaches are likely to be vital against the interconnected challenges of emerging infectious diseases.

## Data Availability

All analysis code will be made available on github (eeg31/detectability), with the exception of data dependencies which can be acquired according to the data policies of their source manuscripts.

## Data availability

All analysis code will be made available on github (eeg31/detectability), with the exception of data dependencies which can be acquired according to the data policies of their source manuscripts ^17–20,24,76,77^.

## Acknowledgements

EEG and EP are supported by the Gates Cambridge Trust [Bill and Melinda Gates Foundation OPP1144]. CJC was supported by a postdoctoral fellowship from the Georgetown Environment Initiative. JLNW and OR are supported by the Alborada Trust. We would also like to thank Dr. Freya Shearer and Dr. Jane Messina for their help accessing and interpreting spatial incidence estimates for endemic diseases, as well as the authors of the studies and datasets on which this work depends.

## Notes

### Competing Interest Statement

The authors have declared no competing interest.

